# Clustering of susceptible individuals within households can drive measles outbreaks: an individual-based model exploration

**DOI:** 10.1101/2019.12.10.19014282

**Authors:** Elise Kuylen, Lander Willem, Jan Broeckhove, Philippe Beutels, Niel Hens

## Abstract

When estimating important measures such as the herd immunity threshold, and the corresponding efforts required to eliminate measles, it is often assumed that susceptible individuals are uniformly distributed throughout populations. However, unvaccinated individuals may be clustered in a variety of ways, including by geographic location, by age, in schools, or in households. Here, we investigate to which extent different levels of within-household clustering of susceptible individuals may impact the risk and persistence of measles outbreaks. To this end, we apply an individual-based model, Stride, to a population of 600,000 individuals, using data from Flanders, Belgium. We compare realistic scenarios regarding the distribution of susceptible individuals within households in terms of their impact on epidemiological measures for outbreak risk and persistence. We find that higher levels of within-household clustering of susceptible individuals increase the risk, size and persistence of measles outbreaks. Ignoring within-household clustering thus leads to underestimations of measles elimination and outbreak mitigation efforts.

## Introduction

The global elimination of measles is an important goal of the WHO^1^. To reach this goal by 2020, the WHO has set the target of 95% vaccination coverage, both at the country and district level. Many regions are close to reaching this target, or have vaccination coverage levels that already exceed this threshold. Nevertheless, outbreaks of measles still occur in these same regions^2–4^.

Increasing attention is being devoted to explain why outbreaks still occur in regions with high vaccination coverage^5–7^. Finding the answer to this question might give us an indication of where the barriers for successful elimination of measles lie and enable us to make the final push for eradication of this disease.

Such an explanation is undoubtedly multifaceted, relating to vaccination coverage and vaccine effectiveness over time, as well as to the intricacies of social interactions between individuals in populations. In relation to the latter, one potentially contributing factor is that susceptible individuals might not be randomly distributed in a population, and that this may influence to which extent an overall high vaccination coverage avoids outbreaks from occurring or persisting.

The traditional formula to estimate a threshold for herd immunity states that a fraction as large as 1 − (1*/R*_0_) should be immune to infection for herd immunity to be ensured. With an R_0_ for measles between 12 and 18, a population immunity level of 95% should thus be enough to stop the disease from spreading^8^. However, this formula is based on the assumption that susceptible individuals are evenly distributed throughout the population and that mixing occurs homogeneously. This is, however, not realistic, and may lead to an underestimation of the herd immunity threshold^9^.

When susceptible individuals are clustered in a population, this may lead to a higher risk for measles outbreaks to occur, even when a high level of population immunity already exists^10,11^. Susceptible individuals in a population may be clustered in a number of ways. One way of clustering, which has already received some attention, and which has turned out to have an impact on the risk for measles outbreaks, is clustering based on geographical proximity^9,12,13^. Such findings support policy and provide a basis to set routine immunization levels not only at country-but also at district level, or to recommend campaigns in under-immunized regions^9,12^.

Another way in which individuals can be clustered is by age. As social contact patterns are highly assortative with age^14^, susceptible individuals belonging to under-immunized age categories are likely to cluster and thus to be at greater risk for measles infection^9,15,16^. Other reasons for the clustering of un-vaccinated individuals can be socio-cultural and/or socio-economic, i.e. related to religion, ideology, educational attainment, income, or psychological receptiveness to rumours of vaccine associated adverse events^2,17–20^. Finally, the household one belongs to can also be influential in determining one’s vaccination status. Parents who have a negative attitude towards vaccination, or no access to vaccination resources, might leave one or more of their children un-vaccinated^21–23^. This can lead to an accumulation of susceptible children and young adults within the same households.

In the current paper, we aim to investigate the influence of within-household clustering, using an individual-based model. In this type of model, the population consists of unique entities that interact with each other under the constraints of a given social structure and transmission model. This makes it possible to model multiple levels of heterogeneity in the population, such as differences in social contact behaviour or in preventive behaviour. Such sources of heterogeneity are especially relevant for diseases - such as measles - for which a high level of immunity already exists within a population^24^.

In Flanders, Belgium, 96.2% of infants were vaccinated with measles-containing vaccines at 1 year of age, and 93.4% of children at 10 years of age, as of 2016^22,25^. This is above - or, for adolescents, close to - the threshold that was set by the WHO. However, this does not mean that Flanders is safe from measles outbreaks. Regions enjoying similarly high overall vaccination coverage have recently experienced severe outbreaks^4,6,26^, and in Flanders, smaller outbreaks regularly occur^2,3,17,27^.

In this paper, we will use Stride, a previously developed individual-based model for the transmission of infectious diseases^28^, to investigate the impact of different levels of within-household clustering of susceptible individuals on the risk for measles outbreaks. To parametrise the model we used data from Flanders, Belgium. We investigate how the level of clustering within households impacts the probability of an outbreak to occur, the distribution of final outbreak sizes, and the distribution of the effective reproduction number.

## Methods

Stride is a stochastic model, meaning that some of its processes, such as the simulation of social contacts and disease transmission have a probabilistic component. The simulator is designed to be versatile: by supplying different input files, it is possible to simulate a broad range of populations, scenarios and diseases. The core logic of the model is implemented in C++, making it highly portable and open to performance-optimization^29^. Stride is an open-source project. Its code is maintained in a public Github repository^30^. We briefly discuss the main features of the model and simulator here, and refer to Kuylen et al. (2017)^28^ for more details.

### Population

The population we used consisted of 600,000 individuals, a sample of 10% of the population in Flanders. The synthetic population consists of reference households, obtained from a social contact survey conducted in Flanders in 2010–2011^31,32^. The geographic distribution of these households is based on 2001 census data^33^. The population is closed, meaning that no births or deaths occur during the simulation. Children are assigned to a daycare (0–2 years old), preschool (3–5 years old), primary (6–11 years old), secondary (12–17 years old) or tertiary (18–23 years old) school based on enrolment statistics for Belgium, as acquired from Eurostat^34^. Adults (18–64 years old) are assigned to a workplace based on age-specific employment data and aggregated workplace size data from Eurostat and commuting data from the 2001 census. To account for general contacts, all individuals are also assigned to two artificial ‘communities’. One of these represents the general contacts made during the week, while the other represents those made during the weekend, as social contact behaviour differs substantially between work week and weekend days. Each of these communities consists of 1,000 individuals on average, which is in line with the size of such communities used in a previous model^35^. The age-dependent contact rates that determine social behaviour in each of these contexts - to which we refer as ‘contact pools’ - are based on data from a social contact survey in Flanders^32,36^. We did not model holidays.

### Immunity

At the initiation of each simulation, each individual is defined to be either immune or susceptible to the simulated infection. Individuals are marked as immune based on their age and the target level of household-based clustering. This target level of clustering is in turn determined by an input parameter with a value between 0 and 1. In short, this parameter represents the probability that if one person - born since 1985 - in a household is vaccinated, all other persons born since 1985 in that household will also be vaccinated.

To immunize the population, we follow the procedures that we describe below. The *Immunizer* class samples individuals from the population to immunize at the beginning of the simulation. It needs two parameters to do this. First, it needs an age-specific distribution of immunity: for each age, we need to know the fraction of immune individuals. The distribution we used is a projection of the age-dependent immunity levels to measles in 2020 in Flanders, Belgium. It is based on projections made by Hens et al.^27^, and was obtained in the same way as was described in a more recent paper^16^. Secondly, the *Immunizer* also needs to be supplied with a target clustering level, as was described above.

When sampling immune individuals in the population, we first sample among those individuals that are 35 years of age or older – i.e. individuals that were born before 1985. As measles virus circulation was not substantially affected by vaccination until the mid 1980s in Belgium, we assume that the majority of individuals born before 1985 acquired natural immunity^37^. As such, we assume that clustering of susceptible individuals within households as a result of vaccination decisions does not yet play a role in this age category. To immunize individuals born before 1985, we use the following procedure. First, we calculate for each age older than 35 how many immune individuals should be in the population, based on the age-specific immunity level and the number of individuals of that age that are present in the population. Next, we select a random individual. If this individual is still susceptible, and there are not yet immune individuals of the age this person belongs to, we make them immune. We repeat this procedure until all age-dependent immunity quota for individuals older than 35 have been fulfilled.

Next, we immunize individuals younger than 35 years of age, taking into account the clustering level that was provided as an input parameter. First, we again calculate the required number of immune individuals per age given the target coverage. Secondly, we draw a random individual from a random household. If there are not yet enough immune individuals in their age-category, and provided they are still susceptible, we set this individual’s health state to ‘immune’. Next, we compare a random draw between 0 and 1 to the clustering level. If the draw is smaller than the clustering level, we immunize all other individuals in this household that are younger than 35 years of age. We do this within a 10% range of the constraints of age-dependent immunity levels. If the random draw is greater or equal to the target clustering level, we go back to step 2, sampling a random individual from a random household. We repeat this procedure until all age-dependent immunity quota have been fulfilled.

### Contact and transmission events

The simulator moves forward in discrete time-steps of one day. Each time-step consists of two phases. First, the health status and corresponding presence in contact pools is updated for each individual. Second, the actual contacts and transmission events within each contact pool are simulated.

We adhere to the following natural history for measles. We draw 4 durations from a distribution for each individual: the duration of the incubation period, the duration of the latent period, the duration of the infectious period, and the duration of the symptomatic period. The duration of the incubation period is sampled from a log-normal distribution with median 12.5 and dispersion 1.25^38^. At the end of the incubation period, individuals become infectious for a duration of 6 to 8 days^39^. Next, after either 2 or 3 days of being infectious but still asymptomatic, individuals become symptomatic^38^. When individuals become symptomatic, it is assumed that they will remain home and thus only make contacts with persons within their own household. This symptomatic period lasts 6 to 8 days, and we assume that each primary infected individual eventually becomes symptomatic^40^.

To simulate contacts and transmissions within each contact pool, we use the age- and context-dependent contact rates described above. When a contact occurs between an infectious individual and a susceptible individual, a transmission probability is used to determine whether an actual transmission of the virus occurs. This P_transmission_ is supplied as an input parameter.

### Simulations

First, we established a relationship between the transmission probability P_transmission_ and R_0_, the basic reproduction number. We did this by running a large number of simulations, for transmission probabilities ranging from 0 to 1. We tested 21 values for the transmission probability, and ran 1,000 stochastic simulations for each of these values. Seeds to initialize our random number generator were generated from a non-deterministic, machine-specific source. We kept track of the number of secondary cases that was caused by one index case in an otherwise completely susceptible population and used this as an estimator for R_0_. We ran each simulation for 30 days, as we were only interested in the transmissions made by the index case, who recovered after – at most – 27 days. From the 21,000 data points we collected in this way, we fitted a function, allowing us to predict a corresponding 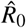 value from a given P_transmission_ value. We fitted the data using the *optimize*.*curve_fit* function in the *scipy* python package^41^. This uses a non-linear least squares method to fit the simulation data to a given function. In our case we used a function of the form *a* + *b* ∗ log(1 + *P*_*transmission*_).

Secondly, we ran simulations over the course of 730 simulated days, until no more new infections were being recorded, to track the full course of the epidemic. We tested 13 values for the transmission probability between 0.2 and 0.8 We also tested 5 different target levels for the clustering of susceptible individuals within households (0.00, 0.25, 0.50, 0.75, 1.00), defined as the probability that another person (aged below 35 years of age) within the same household is immune if one person (aged below 35 years of age) in a household has been vaccinated. For each of these 65 scenarios, we ran 1,000 simulations.

We started each simulation with the introduction of one random infected individual in the population. Before the first time-step, we recorded the number of immune and susceptible individuals in each age category. To determine to which extent susceptible individuals were actually clustered within households, we calculated a ‘household assortativity coefficient’ for the entire population at the beginning of each simulation. To do this we calculated the assortativity based on immunity status (susceptible or immune) in a network of individuals connected by household relations^42,43^. We constructed a network, the nodes of which represented the individuals in the population. Each node has one attribute, ‘susceptible’, which can be either true or false. An edge between two nodes exists if two individuals belong to the same household. We used the networkX python package to construct the network and calculate the attribute assortativity coefficient^44^.

During the simulation, we also keep track of new transmissions. This enables us to determine the total number of cases over the course of the simulation. As we record who infects whom, it is also possible to calculate an effective R - i.e. how many persons the index case infects on average.

To run these simulations, we used the Python environment we created for Stride (PyStride), and used the *multiprocessing* package to run multiple simulations in parallel^45^. We ran all simulations on a Linux machine, using 8 cores (16 with hyper-threading).

## Results

### Relationship P_transmission_ ∼ R_0_

First we established a relationship between the transmission probability P_transmission_ and R_0_. We estimated R_0_ as the expected number of secondary cases one infected individual would infect in an otherwise completely susceptible population.

In Fig. 1, the median (solid blue line), mean (dotted pink line), and 95% percentile interval (grey shape) over 1,000 simulations for each of 21 values of P_transmission_ between 0 and 1 are shown. We see that both the mean and the median number of secondary cases steadily increases as the transmission probability is increased. The shape roughly follows that of a logarithmic function, which is the reason why we chose to fit the data to a function of the form *a* + *b* ∗ log(1 + *P*_*transmission*_). We observe that the 95% percentile interval widens as the transmission probability increases, but that nevertheless, mean and median remain very similar over all values of P_transmission_.

**Figure 1.**
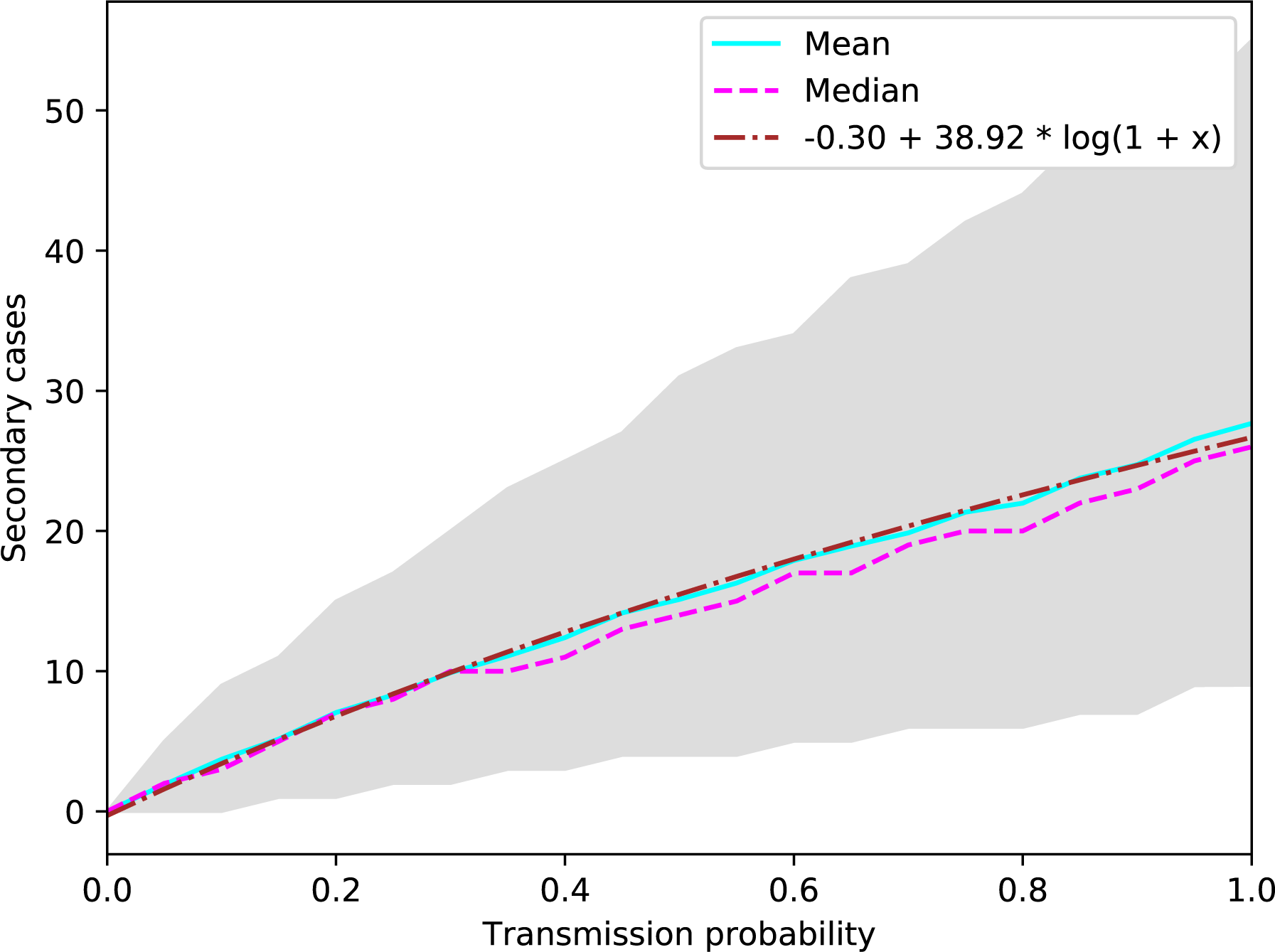
Mean (solid blue line), median (dotted pink line) and 95% percentile interval (grey shape) of secondary cases caused by an index case over 1,000 simulations per value for P_transmission_. The dotted brown line represents the function we fit to the data.

We fitted the data using the *optimize*.*curve_fit* function in the *scipy* python package^41^. This gave us the formula shown in equation (1) to estimate R_0_ for a given P_transmission_.

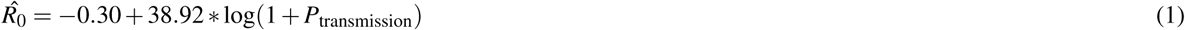

In Fig. 1, we also added the fitted function for comparison (dotted brown line). When P_transmission_ is 0, we observe a corresponding 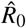 value of 0. We observe that the fitted function closely follows both the mean and median number of secondary cases we observed in our simulations. Based on the formula we found, a realistic range for transmission probabilities for measles in our model is between 0.372 and 0.601, which corresponds to a value of R_0_ between 12.01 and 18.02, which is in line with the basic reproduction number commonly estimated for measles^46^. We ran simulations for P_transmission_ ranging from 0.2 to 0.8, to be certain to include all relevant R_0_ values (R_0_ ∼ 6.80 to 22.58).

### Population immunity profiles

To get an idea of the way in which immunity is distributed over the population in the different scenarios, we investigated the fraction of susceptible individuals by age, and the assortativity of individuals within households based on their immunity status as described in the ‘Methods’ section. The mean fraction of susceptible individuals per year of age for simulations with clustering level 0, 0.25, 0.5, 0.75 and 1 can be seen in Fig. 2. Each data point is the mean over simulations for 13 values for the transmission probability, and with 1,000 simulations per combination of clustering level and transmission probability, i.e. 13,000 simulations per tested clustering level in total. We also added the 95% percentile interval of the target projections we used as input data in Fig. 2 (grey shape). More detailed distributions of susceptibility by age for clustering levels from 0 to 1 can be found in Supplementary Fig. S1.

**Figure 2.**
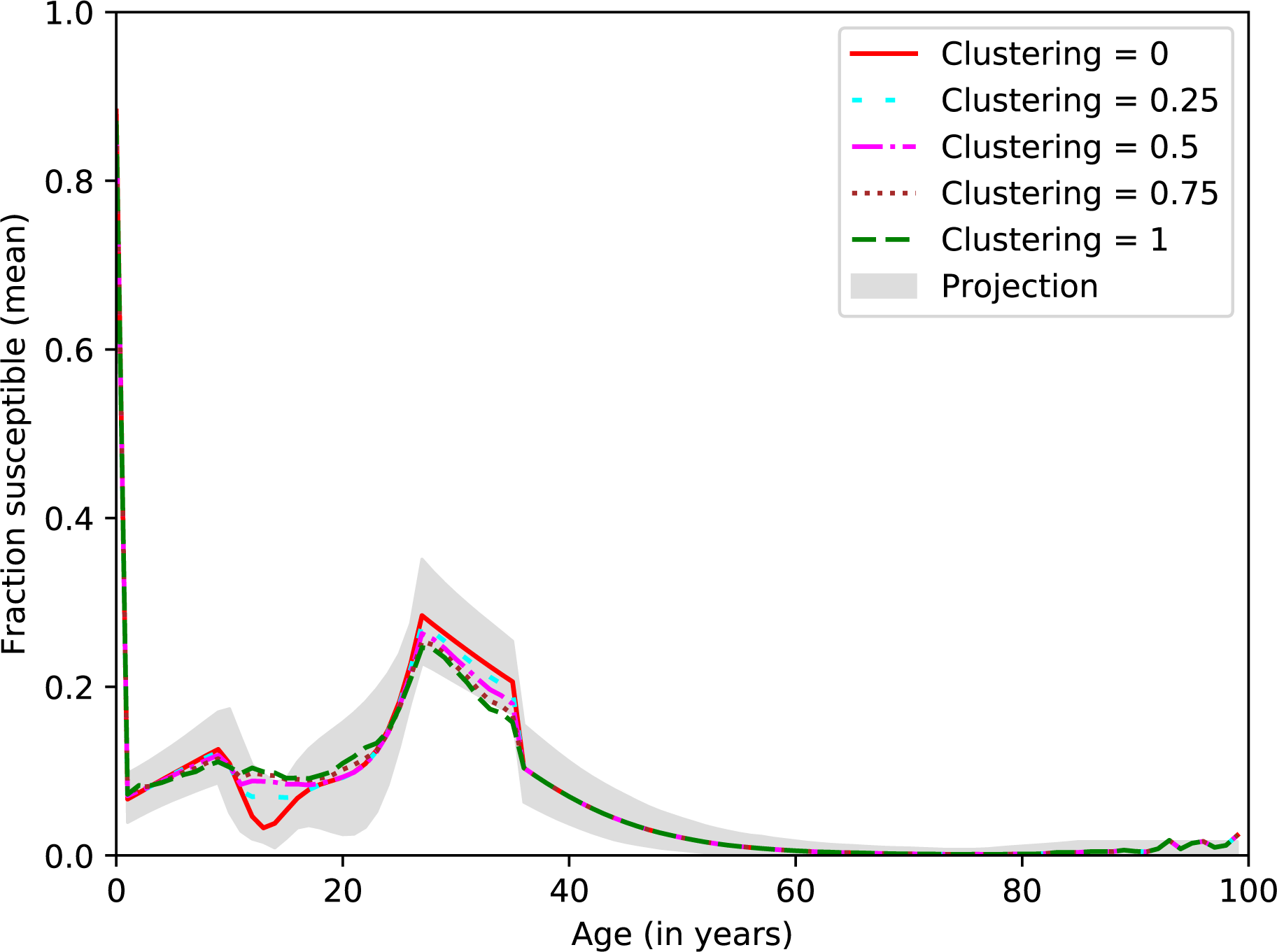
The mean fraction of susceptible individuals per year of age for scenarios with clustering levels 0, 0.25, 0.75 and 1.0. The grey shape represents the 95% percentile interval for the projections we used as input data. Results are based on 13,000 stochastic simulations per clustering level.

We see that overall, the immunity levels in all scenarios seem to be consistent with the target projected immunity levels per age. As the clustering level increases, however, we observe small discrepancies between the simulated and projected immunity levels per age. This can be attributed to the fact that we use a household sample to initialize our population, and that we do not have the ‘right’ household constitutions in our simulated population to accommodate for high levels of clustering within households and age-specific immunity levels at the same time. Slight variations in older age categories can be attributed to fewer individuals being present in these age categories.

We also looked at the relationship between the clustering level we provided as an input parameter to the simulator, and the resulting household assortativity coefficient of the simulated population. The clustering level represents the probability that if one household member under the age of 35 is immune to measles, all other household members under 35 years of age will also be immune. We calculated the household assortativity coefficient – defined as the assortativity of individuals within households, based on immunity status – for clustering levels 0, 0.25, 0.5, 0.75 and 1. We did this for both the entire population and for individuals under the age of 35.

As we increase the clustering level, we observe that the household assortativity coefficient also increases. This is the case when we observe the entire population (Fig. 3a), as well as when we only observe individuals under 35 years of age (Fig. 3b). When we measure the household assortativity coefficient over the entire population, it increases from about 0.3 to 0.5 as the target clustering level increases from 0 to 1. However, when we only take into account individuals aged under 35 years of age, we see an increase from about 0.35 to 0.85 as the clustering level increases from 0 to 1. This discrepancy is as expected, as we only applied household-based clustering of susceptible individuals for individuals age under 35 years of age. Taking into account older individuals when calculating the household assortativity coefficient lowers the coefficient, as these individuals have not been actively clustered by immunity status. We also observe that there is not much variation per clustering level over the different simulations.

**Figure 3.**
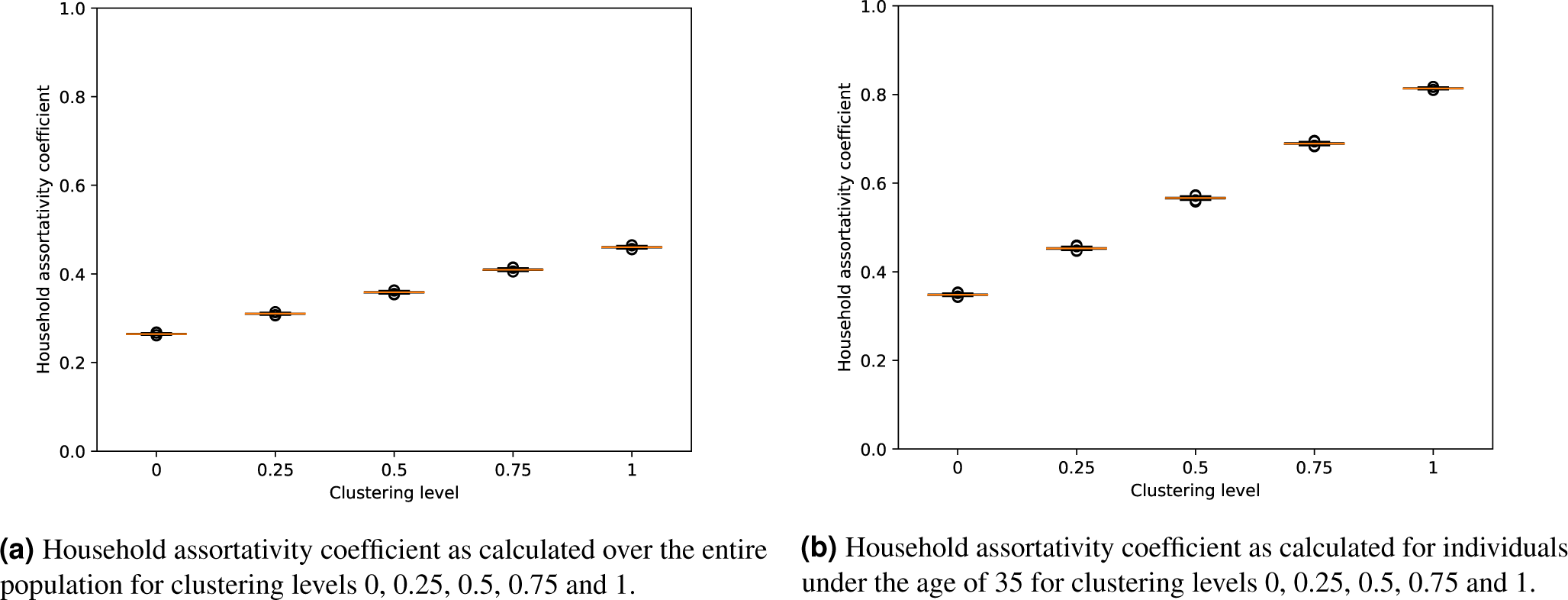
Household assortativity coefficients observed for clustering levels 0, 0.25, 0.5, 0.75, 1. Results are based on 13,000 stochastic simulations per clustering level.

### Effective R

For each combination of transmission probability and clustering level that we tested, we also calculated the effective R. We defined this as the expected number of secondary cases caused by our index case in a partially immune population. In Fig. 4, a heat-map indicates the effective R – as calculated from 1,000 simulations per scenario – for clustering levels 0, 0.25, 0.5, 0.75 and 1 and transmission probabilities between 0.2 and 0.8 (R_0_ ∼ 6.80 to 22.58). We observe that a higher clustering level results in a higher effective R for the same transmission probabilities.

**Figure 4.**
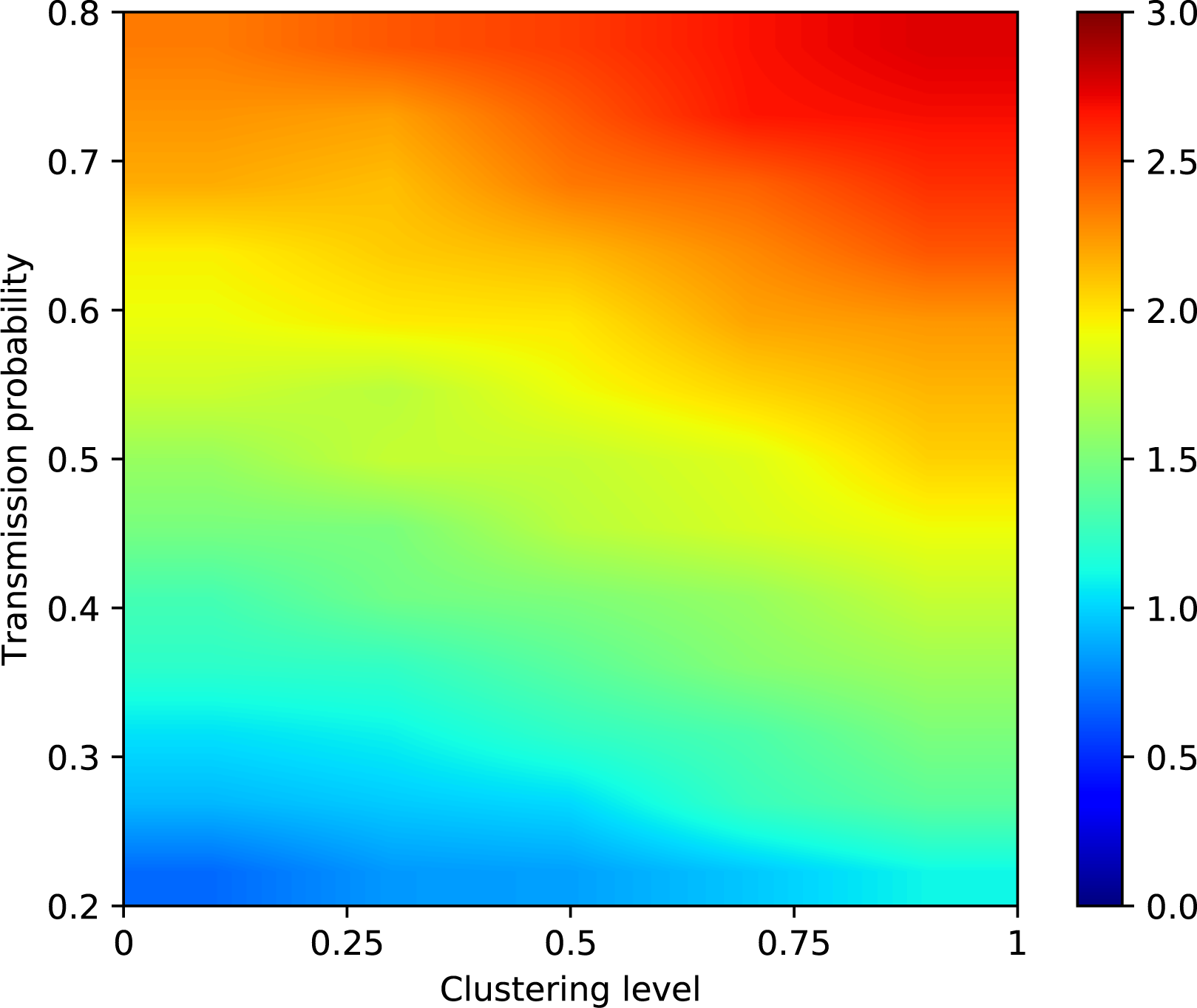
Effective R for clustering levels 0, 0.25, 0.5, 0.75 and 1 and values of P_transmission_ from 0.2 to 0.8 (R_0_ 6.80 to 22.58). Results are based on 1,000 simulations per scenario.

The effect is stronger for lower values of P_transmission_, and becomes weaker as P_transmission_ increases. This is clear when we investigate the percentage the effective R increases when increasing the clustering level from 0 to 1. An overview of the increase in effective R when increasing the clustering level from 0 to 1 can be found in Table 1. We see that, while the increase in effective R is above 60% when P_transmission_ is 0.2 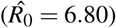, it decreases to about 18% for a transmission probability of 0.6 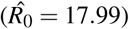.

**Table 1.**
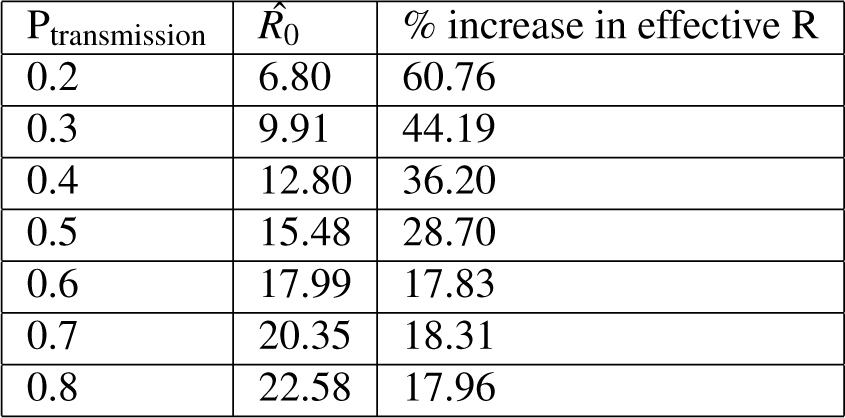
Percentage by which the effective R increases when the clustering level increases from 0 to 1, for different values of PTRANSMISSION. Results are based on 1,000 simulations per combination of P_transmission_ and clustering level.

### Outbreak persistence and size

We are also interested in the number of outbreaks that persist versus the number of outbreaks that goes extinct after only a few cases. To investigate this, we first define what we regard as persistence versus what we regard as extinction. For this purpose, we examined the frequency of outbreak sizes for different values of P_transmission_. Histograms for outbreak sizes – defined as the total number of infected cases after 730 days – over clustering levels 0, 0.25, 0.5, 0.75 and 1 can be seen in Supplementary Fig. S2 (P_transmission_ = 0.20 to 0.35, 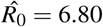 to 11.38), Supplementary Fig. S3 (P_transmission_ = 0.40 to 0.55, 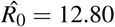 to 16.76), and Supplementary Fig. S4 (P_transmission_ = 0.60 to 0.80, 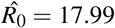 to 22.58). We see that, for all transmission probabilities, outbreaks either die out after only a few cases (bins on the far left), or persist and infect a large part of the susceptible population (bins on the far right). These histograms also show that, as the clustering level increases, outbreaks tend to become larger: outbreaks that persist with clustering level 0 (red bars) are generally smaller than outbreaks that persist with clustering level 1 (green bars). Based on these plots, we set the extinction threshold - above which we will regard an outbreak as persistent - at 5,000 cases.

Based on this threshold, we calculated the fraction of outbreaks that persisted in each scenario. A heat-map showing the probability of persistent outbreaks for clustering levels 0, 0.25, 0.5, 0.75, and 1 and transmission probabilities between 0.2 and 0.8 (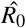 between 6.80 and 22.58) is shown in Fig. 5. With P_transmission_ = 0.2, we observe no persistent outbreaks, regardless of the clustering level. However, with a transmission probability of 0.3, which corresponds to an R_0_ of about 9.91, there are still no persistent outbreaks with a clustering level of 0 or 0.25, but persistent outbreaks start to appear for clustering levels 0.5, 0.75 and 1. For a transmission probability of 0.35 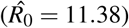, we observe persistent outbreaks for all clustering levels.

**Figure 5.**
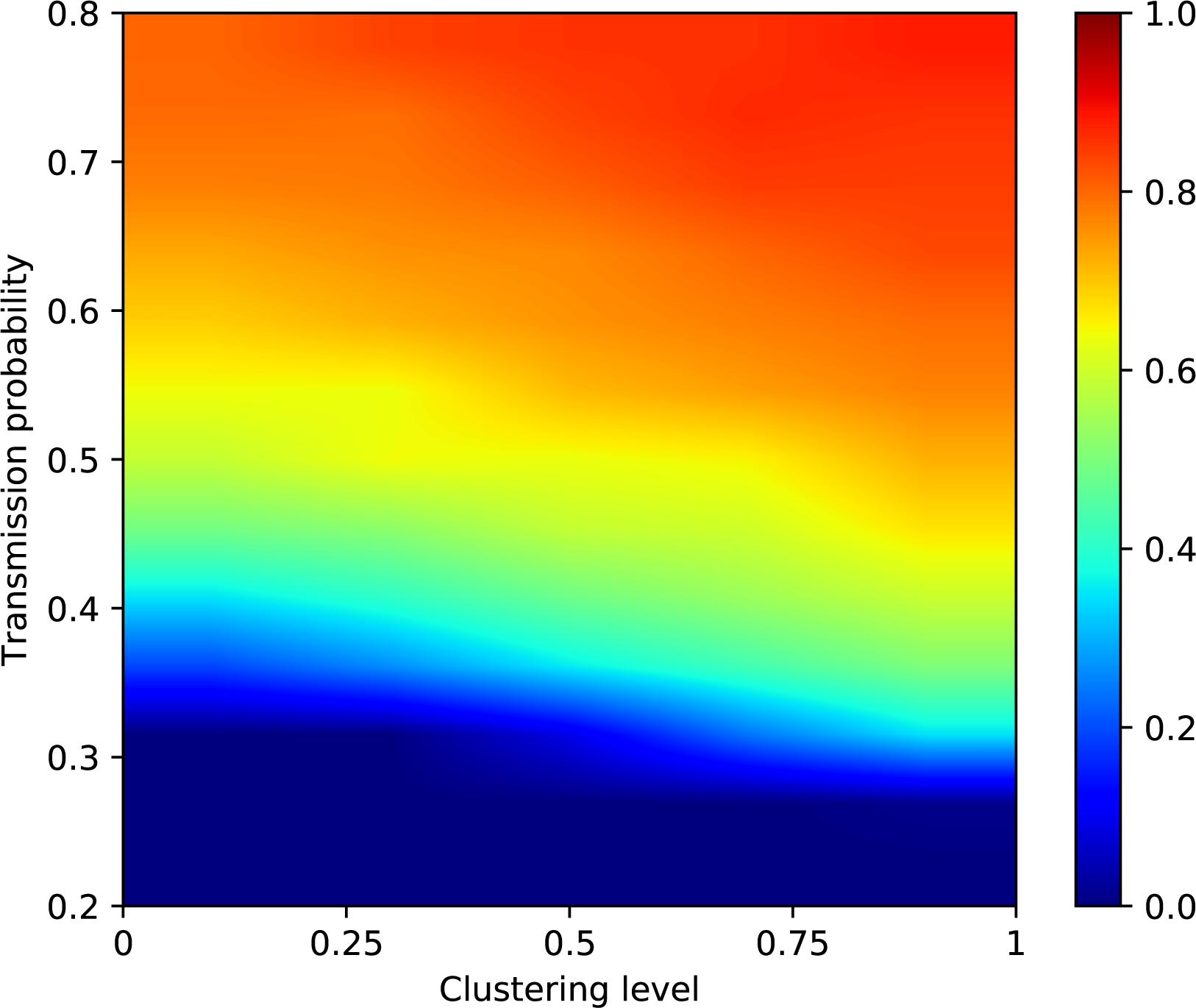
Fraction of persistent outbreaks for clustering levels 0, 0.25, 0.5, 0.75, and 1, with values for P_transmission_ ranging from 0.2 to 0.8. Per scenario, we ran 1,000 simulations. Extinction threshold = 5,000 cases.

However, at a clustering level of 0, only about 20% of outbreaks is persistent, while this increases to about 50% for a clustering level of 1. For larger transmission probabilities, the same trend can be observed: as the clustering level increases, so does the percentage of outbreaks that persist. The effect is stronger for lower values of P_transmission_, and less pronounced for higher values of P_transmission_. A more detailed overview of outbreak probabilities per value of P_transmission_ can be found Supplementary Fig. S5 (P_transmission_ = 0.20 to 0.35, 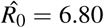 to 11.38), Supplementary Fig. S6 (P_transmission_ = 0.40 to 0.55, 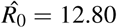 to 16.76), and Supplementary Fig. S7 (P_transmission_ = 0.60 to 0.80, 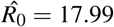 to 22.58).

Similar trends can be observed when we look at the escape probabilities for the different scenarios. We define the escape probability as shown in equation (2), with N_susceptible_ the number of susceptible individuals at the beginning of the simulation, and N_cases_, the number of infected cases after 730 simulated days.

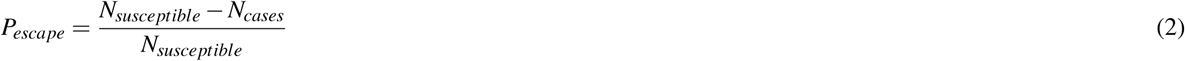

A heat-map for the mean escape probability over 1,000 runs for each scenario (clustering levels ranging from 0 to 1 and transmission probabilities ranging from 0.2 to 0.8 - 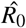 of 6.80 to 22.58 - is shown in Fig. 6. For a transmission probability of 0.2 and 0.25 (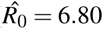 and 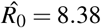, respectively), we see that the escape probability is almost 1 on average for all clustering levels. This is what we would expect, as we observed before that there were no persistent outbreaks at this transmission probability. However, with a transmission probability of 0.3 and 0.35 (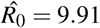 and 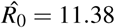 respectively), we see that while the escape probability is still almost one for a clustering level of 0, we observe that the escape probability lowers to about 0.9 for a clustering level of 1. For values of P_transmission_ 0.4 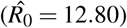 or higher, we see that while the mean escape probability for clustering level 0 is below 1, it decreases further as the clustering level increases. This effect becomes less pronounced for P_transmission_ 0.7 and 0.8 (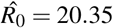 and 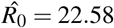 respectively), but it can still be observed. A more detailed overview of the distribution of escape probabilities by clustering level and transmission probability can be found in Supplementary Fig. S8 (P_transmission_ = 0.20 to 0.35, 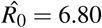 to 11.38), Supplementary Fig. S9 (P_transmission_ = 0.40 to 0.55, 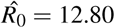 to 16.76), and Supplementary Fig. S10 (P_transmission_ = 0.60 to 0.80, 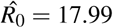 to 22.58).

**Figure 6.**
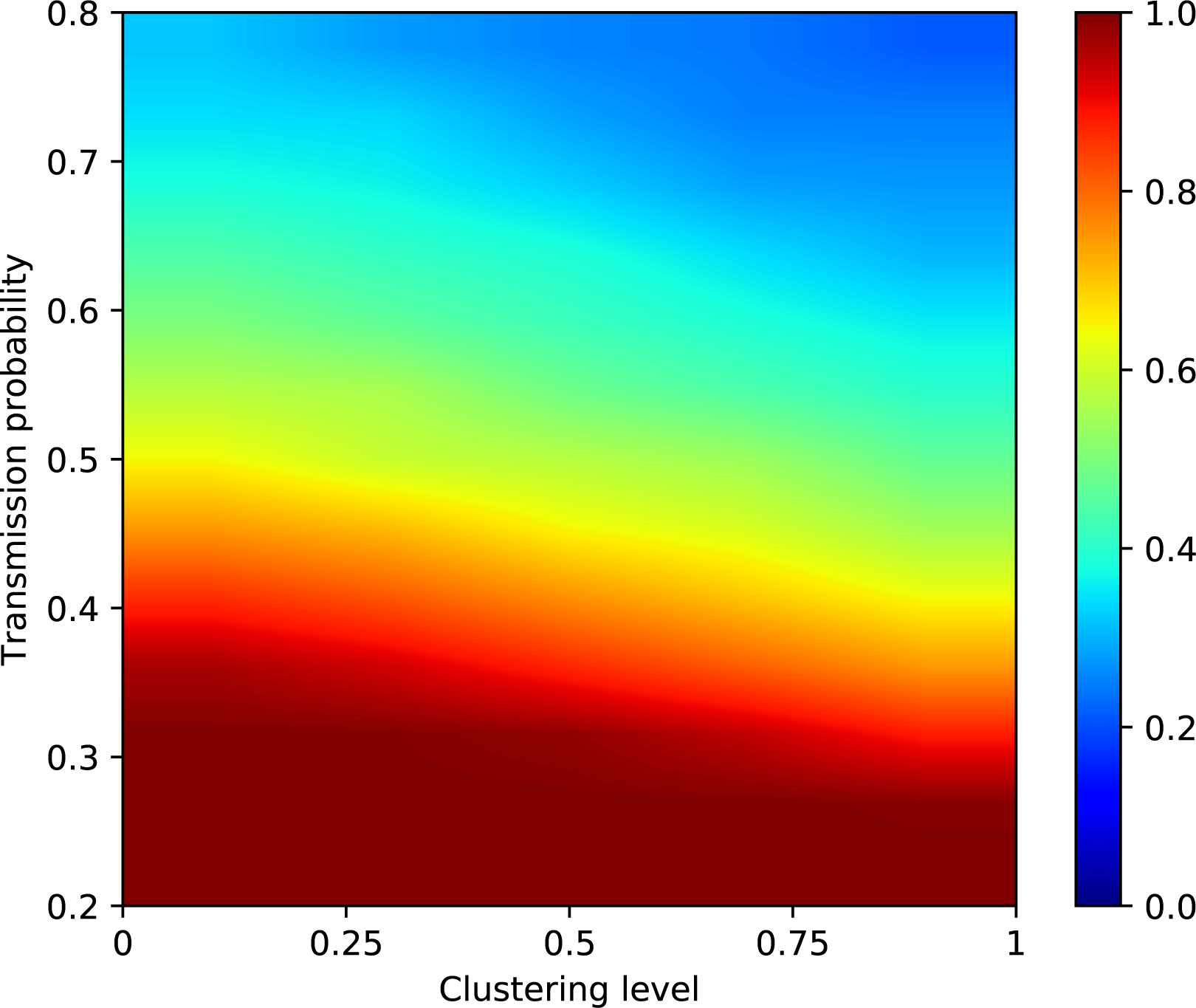
Mean escape probability for clustering levels 0, 0.25, 0.5, 0.75, and 1, with P_transmission_ ranging from 0.2 to 0.8. We calculated the average escape probability over 1,000 runs for each scenario.

**Figure 7.**
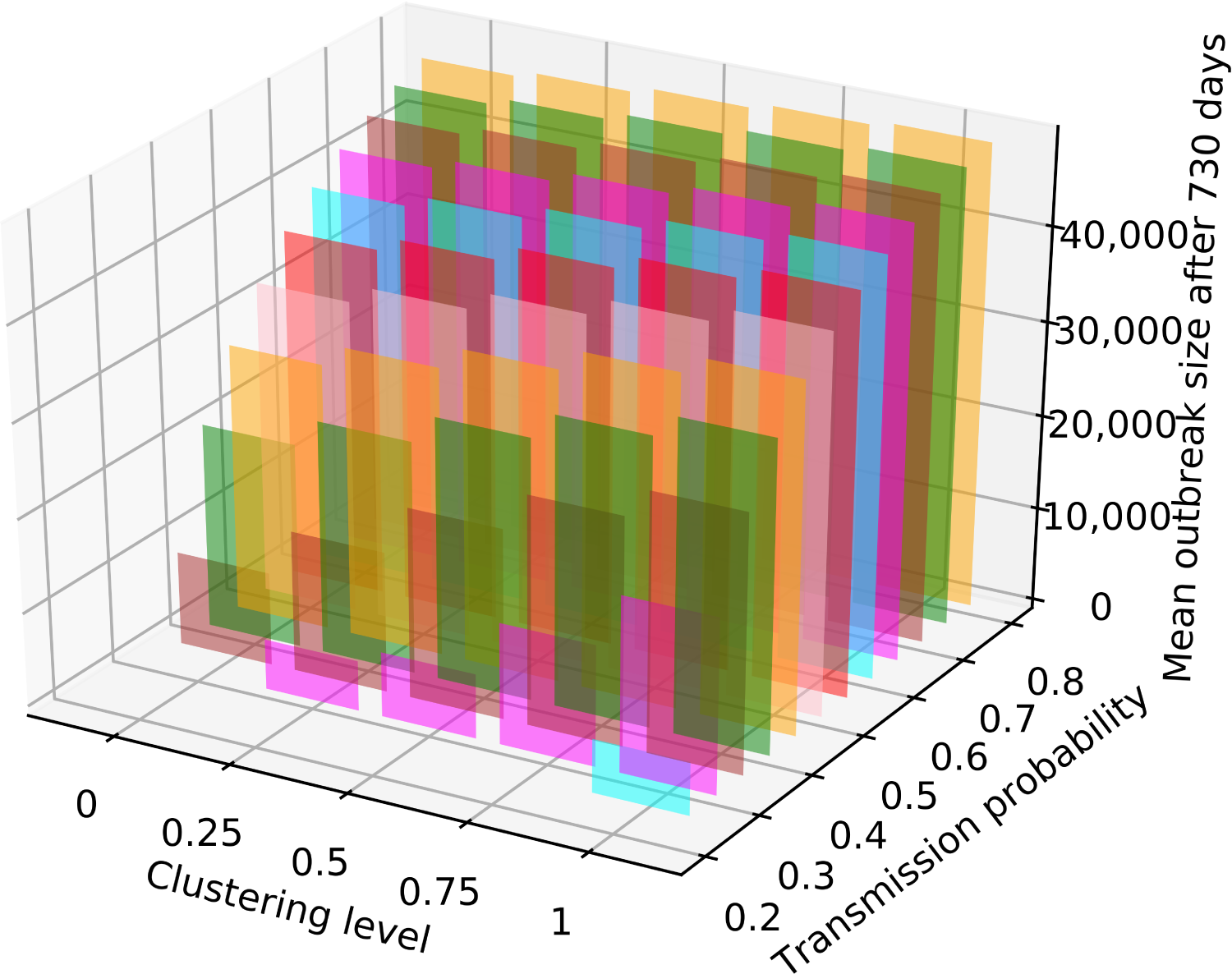
Mean outbreak size (averaged over 1,000 runs per scenario) for persistent outbreaks. Results for transmission probabilities 0.2 to 0.8 (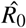 of 6.80 to 22.58) and clustering levels 0 to 1 are shown here. We set the extinction threshold at 5,000 cases.

We also examined the sizes of the simulated outbreaks in the different scenarios. In Fig. **??**, the mean outbreak size for persistent outbreaks over 1,000 runs is shown for clustering levels ranging from 0 to 1 and transmission probabilities ranging from 0.2 to 0.8 (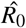 of 6.80 to 22.58). As discussed above, we set the extinction threshold at 5,000 cases.

We observe that the outbreak size when P_transmission_ = 0.2 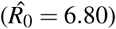 remains 0 at all clustering levels. With a transmission probability of 0.25 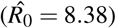, persistent outbreaks only occur at a clustering level of 1. On average, these outbreaks only just reach the extinction threshold of 5,000 cases. For a transmission probability of 0.3 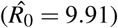, persistent outbreaks occur for clustering levels 0.25, 0.50, 0.75 and 1. With a transmission probability above 0.35 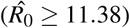, we observe that as the clustering level increases, the average outbreak size also increases. The increase in average outbreak size from clustering level 0 to 1 is steep for transmission probabilities 0.35 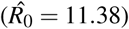 to 0.55 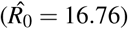. For higher values of P_transmission_, the same trend can be observed, but the increase in outbreak size when the clustering level is increased from 0 to 1 is smaller.

A more detailed representation of the distribution of outbreak sizes by clustering level and transmission probability can be seen in Supplementary Fig. S11 (P_transmission_ = 0.20 to 0.35, 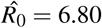 to 11.38), Supplementary Fig. S12 (P_transmission_ = 0.40 to 0.55, 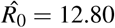 to 16.76), and Supplementary Fig. S13 (P_transmission_ = 0.60 to 0.80, 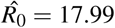 to 22.58). Again, we only looked at persistent outbreaks, and set the extinction threshold at 5,000 cases. As the clustering level increases from 0 to 1, we observe that for all transmission probabilities, the outbreak sizes also increase. This is, again, more pronounced for lower values of P_transmission_. For a transmission probability of 0.35 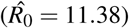, we see that the median outbreak size increases from about 10,000 to about 25,000. For this same transmission probability we notice that there is a higher variance for the outbreak size at lower clustering levels than can be observed at higher levels of clustering. The same can still be observed - albeit to a lesser extent - when we look at results for P_transmission_ = 0.4 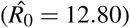. Here the median outbreak size increases from a little over 20,000 to about 35,000 as the clustering level increases from 0 to 1. For higher values of P_transmission_, the median outbreak size also increases as the clustering level increases. However, the variance in outbreak sizes is small for all clustering levels. We also observe that, as P_transmission_ increases, the relative increase in outbreak size becomes smaller as the clustering level increases from 0 to 1.

Finally, we also wanted to know which individuals in the population had the most risk of being infected. For each scenario, we calculated the mean age of infected individuals. As expected, there is not much difference between the different scenarios. With a higher clustering level, infected individuals tend to be slightly younger on average, but are still very close in age to infected individuals when testing a lower clustering level. A heat-map of average ages of infected individuals for transmission probabilities 0.2 to 0.8 (R_0_ ∼ 6.80 to 22.58) and clustering levels 0 to 1 can be found in Supplementary Fig. S14.

## Discussion

We looked at the impact of different levels of within-household clustering of susceptible individuals on the risk for measles outbreaks. We did this by including age-dependent immunity rates and household-based clustering into Stride, an individual-based simulator for the transmission of infectious diseases.

We established a measure for estimating the level of household-based clustering of susceptible individuals in a population: the household assortativity coefficient. We calculated this measure as the assortativity based on immunity status within a network of nodes – representing individuals – connected by household relations – meaning that two nodes were connected if they belonged to the same household.

We also simulated outbreaks of measles by introducing one infected individual into the simulated population of Flanders, Belgium and following the evolution of the outbreak for 730 simulated days. We found that the level of within-household clustering of susceptible individuals has an impact on important measures for outbreak risk. The effective R increases as the clustering level increases. This is true for all tested transmission probabilities (0.2 to 0.8, which we calculated to be equivalent to an R_0_ of about 6.80 to 22.58). However, the relative increase in effective R is higher for lower values of P_transmission_ and is less pronounced for higher transmission probabilities.

This means that, as more clustering of susceptible individuals within households occurs in a population, an index case will on average infect more cases compared to a population with a lower household assortativity coefficient. As such, household-based clustering of susceptible individuals has an impact from the very beginning of an outbreak.

It also plays a role in the rest of the evolution of measles outbreaks. We observed that outbreaks have a higher probability of persisting when there is a higher level of household-based clustering of susceptible individuals. When we set the threshold for persistent outbreaks at 5000 cases, we saw that while there were no persistent outbreaks with a transmission probability of 0.3 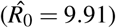 and a clustering level of 0 and 0.25, bt persistent outbreaks did appear when the clustering level was at 0.5, 0.75 or 1. For higher values of P_transmission_, we noticed that the probability for an outbreak to persist increased as the clustering level increased.

We observed the same trend when looking at escape probabilities: as the clustering level increases, the escape probability decreases, meaning that a larger fraction of susceptible individuals will get infected over the course of an outbreak. This can also be seen when looking at the sizes of persistent outbreaks. On average, outbreaks become larger when the level of household-based clustering increases. As such, a higher level of clustering of susceptible individuals within households entails a higher risk for susceptible individuals to become infected whenever an infected individual is introduced in the population.

These effects are strongest for transmission probabilities between 0.3 and 0.6 (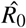 between 9.91 and 17.99). For larger values of P_transmission_, the effects of increasing the level of household-based clustering of susceptible individuals become less pronounced, but can still be observed.

We conclude that, as the level of household-based clustering of susceptible individuals has an impact on the risk for measles outbreaks, it is important to include this form of clustering of susceptibility into models for measles transmission in highly vaccinated populations. Omitting this form of clustering would lead to an underestimation of the risk for outbreaks and thus of the efforts needed to inhibit the spread of measles and to achieve measles elimination. Taking into account within-household clustering, and potentially other ways in which susceptible individuals can be clustered in a population, provides new and more accurate herd immunity thresholds for measles.

Some limitations need to be taken into account when interpreting these results. Firstly, the population we used in Stride is closed. No individuals are born or die over the course of the simulation. However, for a childhood disease such as measles, long-term simulations would benefit from having a demographic component.

Furthermore, the projections of age-specific immunity levels in 2020 we used as input for our simulations were partially based on a serological survey^25^. The serological survey information is based on antibody titre levels, which is a humoral immune response indicator. However, cellular immune response mechanisms – which are only partially correlated with humoral response markers – can still provide immunity even when antibody levels are low or undetectable^47^. The levels of immunity that we have used may thus represent an underestimation of the actual immunity levels against measles in Flanders.

On the other hand, even when an individual’s humoral immunity levels are considered protective, exposure studies suggest that a sufficiently high infecting dose can still trigger an active infection^47^. This could further increase the importance of within-household clustering of susceptible or partially vaccinated people, as the infecting dose is likely highest for people living in the same household as an infectious individual.

In our simulations, we applied household-based clustering for individuals born since 1985. We assumed that most individuals born before that date had acquired natural immunity against measles. However, in vaccination guidelines, 1970 is used as a cut-off for natural immunity, assuming individuals born before 1970 to have natural immunity against measles^48^.

For this study, we have also made some assumptions in our model of the natural history of measles disease. We assumed that every infected individual will eventually become symptomatic, and that individuals, while symptomatic, will only contact members of their own household. We also assumed that all susceptible individuals had the same chance of becoming infected, and subsequently infectious and symptomatic. We did not take into account variation in the probability and duration of symptoms of previously vaccinated versus unvaccinated susceptible individuals^49,50^. Finally, we also assumed that the durations of the incubation period, latent period, infectious period and symptomatic period were independent from each other within each individual. Future analyses should aim to relax these assumptions.

Additionally, data from observational studies on levels of clustering of susceptible individuals within households should be obtained and used as an input to models for measles transmission. Furthermore, the concept of clustering can be further expanded by examining other types of clustering, including geographical clustering and within-school clustering of susceptible individuals^19^.

Finally, if the underlying reasons for non- or incomplete vaccination can be uncovered and quantified, other relevant types of clustering may be defined in future models. As these models become more realistic, they can be used more effectively to inform policy on measles elimination.

## Data Availability

The datasets generated and analysed during the current study are available from the corresponding author on reasonable request.

## Acknowledgements

EK, LW and PB acknowledge support of the Antwerp Study Centre for Infectious Diseases (ASCID) at the University of Antwerp, and the Research Foundation Flanders (FWO) (research project G043815N and a postdoctoral fellowship 1234620N (LW)). NH acknowledges funding received from the European Research Council (ERC) under the European Union’s Horizon 2020 research and innovation programme (grant agreement 682540 — TransMID).

## Author contributions statement

J.B. and N.H. initiated the research. L.W. provided the input data necessary for the simulations. E.K. designed the experiments, ran the simulations and wrote the main manuscript text. E.K., L.W., P.B., and N.H. interpreted the data generated by the simulations. All authors reviewed the manuscript.

